# Comparison of LIAISON QuantiFERON-TB Gold Plus with QuantiFERON-TB Gold Plus

**DOI:** 10.1101/2022.11.21.22282591

**Authors:** Kamran Kadkhoda, Matthew Dee, John Simeone

## Abstract

With the large global burden of latent tuberculosis (TB) infection (LTBI), screening for LTBI for various reasons has become standard practice including the low-prevalence settings.

The results showed acceptable intra- and inter-run precision as well as cutoff verification across the two analyzers at the afore-mentioned levels (Table 1). Overall, the LIAISON QFT-Plus showed better inter- and intra-run precision compared with QFT-Plus when comparing low-positive TB1/2 samples; however, coefficient of variation (CVs) less than 20% are generally acceptable for any automated test. Cutoff verification showed CVs for QFT-Plus TB1 and TB2, QFT-Plus LIAISON TB1 and TB2 of 5.2%, 4.1%, 5.2%, 5.4%, respectively, demonstrating robustness around the cutoffs.

Altogether, the two methods were highly comparable. LIAISON QFT-Plus is a suitable alternative to batch testing (e.g., using DSX) in high-volume laboratories.

---

With the large global burden of latent tuberculosis (TB) infection (LTBI), screening for LTBI for various reasons has become standard practice including the low-prevalence settings (1, 2). Interferon-gamma release assays (IGRAs) are usually preferred over tuberculin skin test given the limitations of the latter (3). Several versions of IGRAs have been introduced to the global market, each time with an improvement including performance characteristics and laboratory operations (4). We previously published an operational feasibility study using the latest version of IGRAs that is commercially available, LIAISON QuantiFERON-TB Gold Plus (DiaSorin Inc.) (LIAISON QFT-Plus)(5). As a sequel to our previous study, we set out to assess the performance of LIAISON QFT-Plus, including method comparison, intra- and inter-precision and cutoff verification in comparison with the previously-used reference method, QuantiFERON-TB Gold Plus (Qiagen) (QFT-Plus). LIAISON QFT-Plus is based on chemiluminescence immunoassay whereas QFT-Plus is based on enzyme immunoassay, and our laboratory did the test on DSX (Dynex technologies) batch analyzers. To this end, 190 randomly-selected de-identified residual plasma samples were used for the comparisons. Of these stored samples, 97, 23, and 70 had initial negative, indeterminate, and positive results, respectively. For the precision and cutoff verification studies, we compared both TB1 and TB2 result components for which we arbitrarily chose low (0.35-1.0 IU/ml) and high (>1.0 IU/ml) levels. For the cutoff verification, a sample was used with initial values within 20% of the positivity cutoff (0.35 IU/ml) and tested 10 times in one run.

**Table 1.** Intra- and inter-run precision between the reference (QFT-Plus) vs. the test (LIAISON QFT-Plus) methods. For inter-run precision, the sample was run for seven consecutive days. For intra-run accuracy, the same sample was run ten times in one run. CVs were calculated for each repeat. Analyses were performed by the EP Evaluator software (Data Innovations).

| Intra-run precision | Low TB1 | Low TB2 | High TB1 | High TB2 |
| --- | --- | --- | --- | --- |
| *DSX* | 9.0% | 7.6% | 0.0% | 0.0% |
| *LIAISON XL* | 6.4% | 9.0% | 0.0% | 0.0% |
| Inter-run precision |  |  |  |  |
| *DSX* | 15% | 15.9% | 0.0% | 0.0% |
| *LIAISON XL* | 7.3% | 7.3% | 0.0% | 0.0% |

Lastly, the method comparison showed excellent results comparing the two kits/platforms (Table 2). In our study, all 20 indeterminate samples that matched between the two methods had high Nil (>8 IU/ml) levels. The three discordant samples with indeterminate results using QFT-Plus on DSX instrument tested negative on LIAISON QFT-Plus. All three samples had had mitogen responses less than 0.5 IU/ml and very low Nil values and negative TB1 and TB2 results, however, mitogen responses tested all >0.5 IU/ml using LIAISON QFT-Plus. Two of the three discordant results were slightly over the acceptable mitogen cutoff (0.5 IU/ml), which could be due to the potential higher sensitivity of LIAISON QFT-Plus. The third discordant sample with a relatively strong mitogen response suggests that the <0.5 IU/ml result on QFT-Plus was possibly due to an aspiration error which is not uncommon with the DSX instrument. Suboptimal indeterminate agreement is generally not unexpected; however, indeterminate IGRA results typically pose management dilemmas.

**Table 2.** Method comparison between the reference (QFT-Plus) vs. the test (LIAISON QFT-Plus). McNemar test for symmetry passed (*p*=0.250 where *p*<0.05 was considered statistically significant) showing no difference between the two methods. Cohen’s kappa was 97.3% (95% CI: 94.2-100.3%) showing a very high agreement between the two methods. The three discordant samples with indeterminate results using QFT-Plus on DSX instrument tested negative on LIAISON QFT-Plus. All three samples had had mitogen responses less than 0.5 IU/ml and very low Nil values and negative TB1 and TB2 results, however, all mitogen responses tested >0.5 IU/ml using the LIAISON QFT-Plus (0.62, 0.99, and 6.97 IU/ml). Analyses were performed by the EP Evaluator software (Data Innovations).

| QFT-Plus (reference) vs. LIAISON QFT-Plus (test) |  |
| --- | --- |
| Negative agreement | 100% (97/97) |
| Indeterminate agreement | 86.9% (20/23) |
| Positive agreement | 100% (70/70) |
| Overall agreement | 98.4% (187/190) (95% CI: 95.5-99.5%) |

Altogether, the two methods were highly comparable. LIAISON QFT-Plus is a suitable alternative to batch testing (*e*.*g*., using DSX) in high-volume laboratories.

## Data Availability

Available

## Disclaimer

This study was deemed a quality improvement project by the Cleveland Clinic Foundation institutional review board.

## Acknowledgement

The study was sponsored by DiaSorin Inc.

